# Impact of sputum quality on Xpert MTB/RIF Ultra test results for tuberculosis: A multi-country study

**DOI:** 10.64898/2026.04.01.26350003

**Authors:** Caitlin A. Moe, Sraoshi Barua, Shrivaas Vijayan, Alfred O. Andama, John Bimba, Devasahayam J Christopher, Van Luong Dinh, Ha Phan, Grant Theron, William Worodria, Charles Yu, Kristin Kremer, Payam Nahid, Seda Yerlikaya, Claudia M. Denkinger, Adithya Cattamanchi, Monde Muyoyeta

**Author notes:** Contributed equally as first authors (Caitlin A. Moe, Sraoshi Barua). Contributed equally as senior authors (Adithya Cattamanchi, Monde Muyoyeta).

## Abstract

**Rationale:** Sputum-based testing using Xpert MTB/RIF Ultra (Xpert) is the most common molecular testing method for diagnosing tuberculosis (TB).

**Objectives:** To evaluate whether sputum quality influences Xpert positivity and diagnostic accuracy.

**Methods:** We screened consecutive people for presumptive TB in India, the Philippines, Vietnam, Nigeria, South Africa, Uganda, and Zambia as part of the R2D2 TB Network and ADAPT studies. Participants provided 2-3 sputum samples for Xpert and culture reference testing. The quality of the first sputum sample was graded following standardized procedures by trained research staff and used for Xpert testing. We performed logistic regression to evaluate whether sputum grade was independently associated with Xpert positivity, and calculated sensitivity and specificity of Xpert against a culture-based microbiological reference standard (MRS).

**Measurements and Main Results:** Among 1,855 participants, 798 (43%) were female, 348 (19%) were living with HIV (PLHIV), and 1795 (97%) had a cough of ≥2 weeks. Overall, 313 (17%) had a positive Xpert result. Most sputum samples were salivary (83%). Xpert positivity was lowest among salivary samples (16.1%) and highest among purulent samples (31.2%). After adjusting for demographic and clinical variables, there was no significant association between any sputum grade and Xpert positivity. Xpert sensitivity (salivary: 89%, mucoid: 91%, mucopurulent: 87%, purulent: 100%) and specificity (>98%) were high across sputum grades.

**Conclusions:** Sputum quality was not independently associated with Xpert positivity and Xpert sensitivity was high across all sputum grades. These findings support molecular testing of all sputum samples for TB diagnosis regardless of macroscopic appearance.

**Scientific Knowledge on the Subject:** Xpert MTB/RIF Ultra (Xpert Ultra) is a WHO-recommended initial diagnostic test for pulmonary tuberculosis. Historical laboratory practice, derived from the smear microscopy era, has emphasized sputum macroscopic quality, with many programs recommending rejection of salivary sputum as inadequate. Prior studies evaluating sputum quality and Xpert performance yielded mixed results, and were limited by small sample sizes, single-country designs, heterogeneous sputum grading systems, and predominant use of the first-generation Xpert MTB/RIF assay.

**What This Study Adds to the Field:** In this large and geographically diverse evaluation of 1,855 participants across seven high-burden countries, macroscopic sputum quality was not independently associated with Xpert Ultra positivity or diagnostic accuracy. Xpert Ultra demonstrated high sensitivity and specificity across all sputum grades, including salivary specimens, which comprised over 80% of samples. These findings, generated by using a standardized sputum grading protocol and a culture-based microbiological reference standard, provide robust evidence that sputum quality-based rejection criteria are unnecessary for Xpert Ultra testing. Revising laboratory guidelines to accept all expectorated sputum regardless of appearance could improve case detection and reduce diagnostic delays in high-burden settings.

## INTRODUCTION

Tuberculosis (TB) remains the leading cause of death from an infectious agent globally, with an estimated 10 million cases and over 1 million deaths annually.^1^ Accurate diagnosis relies heavily on sputum-based molecular testing. Xpert MTB/RIF Ultra (Xpert Ultra), recommended by the World Health Organization (WHO) as an initial diagnostic test for pulmonary TB, is now widely implemented in high-burden settings.^2^ Xpert Ultra is a nucleic acid amplification test designed to detect both *Mycobacterium tuberculosis* complex (MTBC) and resistance to rifampin (RIF). Xpert Ultra provides improved analytical sensitivity compared to the original Xpert MTB/RIF assay through dual-target amplification and semi-nested PCR. Its rapid turnaround time and minimal operational training requirements make it particularly valuable in high TB burden settings.^3^

Historically, laboratory practice has emphasized sputum macroscopic quality, based on evidence from the smear microscopy era showing that mucoid and purulent sputum correlated with higher bacillary load and smear positivity than salivary or watery sputum.^4,5^ Many national TB programs and laboratory manuals still classify thin or salivary sputum as ‘poor quality’, and recommend rejecting these and requesting repeat samples.^6,7^ However, rejection of salivary sputum may exclude groups already facing diagnostic barriers, such as individuals unable to produce high quality expectorated sputum, including people living with HIV (PLHIV), children, and individuals with early disease.^8,9^ Testing all sputum regardless of macroscopic appearance may enhance early case detection, simplify health facility operations, and improve access to diagnosis, if supported by empirical evidence.

Previous research evaluating sputum quality and Xpert performance yielded mixed and often contradictory findings. Studies using the first generation Xpert MTB/RIF assay in Eswatini, Fiji, and Kenya reported slightly higher positivity or sensitivity in mucoid and purulent specimens.^10–12^ In contrast, studies in Brazil, Botswana, Ethiopia, Uganda, Vietnam, and Egypt observed similar or even higher positivity in salivary sputum.^13–18^ However, these studies applied heterogenous sputum classification schemes and most were small, single-country analyses conducted prior to widespread use of the current generation Xpert Ultra assay.

We therefore conducted a multi-country evaluation to determine whether macroscopic sputum appearance influences Xpert Ultra positivity and diagnostic accuracy. Using a standardized sputum grading protocol across all testing sites, we assessed 1) whether sputum quality is independently associated with positive Xpert Ultra results and 2) whether Xpert Ultra sensitivity and specificity for pulmonary TB vary by sputum quality.

## METHODS

### Study Design and Participants

From June 2024 to March 2025, a multi-country cross-sectional study was conducted within the Rapid Research in Diagnostics Development for TB Network (R2D2 TB Network) and the Assessing Diagnostics At Point-of-Care for Tuberculosis (ADAPT) studies. Consecutive people with presumptive TB at clinics in seven high-burden TB countries (India, the Philippines, Vietnam, Nigeria, South Africa, Uganda, and Zambia) were screened and enrolled as part of studies evaluating novel TB tests.

People were eligible if they were aged 12 years or older and were presumed to have pulmonary tuberculosis based on (1) having cough for ≥2 weeks, or (2) at least one risk factor for TB (HIV infection, history of mining, TB contact) along with a positive screening test result (abnormal chest X-ray or, for PLHIV, C-reactive protein [CRP]>5mg/dL). Participants were excluded if they had undergone preventive or active TB treatment in the past 12 months, taken any anti-mycobacterial medication in the past two weeks, and/or were unwilling to provide informed consent. All study participants provided written informed consent. For this analysis, we included participants enrolled between June 2024 and March 2025 (when sputum grading was performed) and excluded participants for whom sputum induction was required.

### Procedures

Demographic and clinical characteristics were collected from all participants using a standardized form. All participants were asked to provide two or three expectorated sputum samples for Xpert Ultra testing and mycobacterial culture.

### Sputum Quality Grades

After collection or at the time of receipt at the laboratory, trained research staff assessed the sputum quality and assigned a grade to the sputum sample undergoing Xpert Ultra testing following a standardized approach. Staff assessed whether or not sputum samples were blood-stained and then categorized sputum quality in six grades: i) salivary samples (thin or watery appearance with no viscosity and clear in color); ii) mucoid samples (viscous and clear or white in color); iii) purulent samples (thick or stringy and opaque in color with a yellow or green tinge); iv) mucosalivary (any mix of saliva and mucus) v) mucopurulent (any mix of mucus and purulent material); and vi) salivary-purulent (any mix of saliva and purulent material). For analysis, mucosalivary and salivary-purulent grades were collapsed into the salivary grade, leaving four sputum quality grades for analysis. To promote consistency across study sites, research staff received standardized training on sputum grading using reference images and written definitions (see Supplement). Periodic refresher training and monitoring were conducted to ensure adherence to the grading protocol across participating laboratories.

### Sputum-based Xpert Ultra testing

Sputum was tested with Xpert Ultra according to the manufacturer’s instructions. If the initial test result was non-actionable (*i.e*., error, invalid or no result) or “Trace” positive, then Xpert Ultra testing was repeated. Participants were classified as Xpert-positive if the initial sputum Xpert Ultra result was positive with a semi-quantitative grade of Very Low or higher. If the initial result was trace positive, then participants were classified as Xpert-positive if the repeat test was also positive (any grade) or if the sputum culture result was positive.

### Reference standard

For diagnostic accuracy analyses, we used a microbiological reference standard (MRS) based on sputum culture results. Two sputum specimens were cultured in liquid media for each participant in accordance with each study site’s standard mycobacteriology laboratory protocols. Participants were classified as TB-positive if one or more culture results were positive for MTBC. Participants were classified as TB-negative if both culture results were negative. TB status was considered to be indeterminate if both cultures were contaminated or one culture was contaminated and the other negative.

### Data analysis

Xpert Ultra positivity was calculated overall and for each sputum grade. Blood-stained sputum was separately compared to non-blood stained sputum, as sputum grade and bloodiness were assessed separately. A Pearson chi-square test was performed to evaluate whether the distribution of Xpert Ultra semi-quantitative grades differed between salivary vs. high quality (defined as mucoid, mucopurulent, or purulent) sputum. Logistic regression models were constructed to assess whether sputum quality is independently associated with Xpert Ultra positivity. Bivariate models were constructed for sputum grade and Xpert Ultra positivity, and variables associated with both at the significance level of p < 0.2 were included in the adjusted model. Sensitivity and specificity were estimated overall and for each sputum grade against the MRS. Participants with non-actionable Xpert Ultra results or indeterminate MRS results were excluded. Differences in sensitivity and specificity were compared by sputum grade using Poisson regression models adjusted for the same covariates as in the primary logistic regression model. In sensitivity analyses and robustness checks, associations between sputum grade and continuous variables (cycle threshold [Ct] values, time to culture positivity) were assessed using linear regression factors associated with salivary vs. non-salivary sputum were assessed using logistic regression. Analyses were conducted in R version 2025.05.0+496 and Stata version 18.

### Ethics statement

This study was registered on ClinicalTrials.gov (NCT04923958 and NCT05941052). Written informed consent was obtained from all study participants. Ethical approval for the study was obtained from research ethics committees at the University of California San Francisco (USA), the University of Heidelberg (Germany), Christian Medical College Vellore (India), KNCV Tuberculosis Foundation (the Netherlands), the Abuja Health Research Ethics Committee (Nigeria), De La Salle Health Sciences Institute (Philippines), Stellenbosch University (South Africa), Makerere University (Uganda), Ministry of Health Ethical Committee in National Biological Medical Research (Vietnam) and the University of Zambia (Zambia).

This manuscript adheres to the STARD guidelines for reporting diagnostic accuracy studies.^19^

## RESULTS

### Participant characteristics

Of 2,441 participants screened for enrollment from July 2024 to March 2025, 319 (13.1%) were excluded (**Figure 1**). Of 2,122 participants enrolled, 218 who required sputum induction and 49 who had a missing sputum grade were excluded from the analysis.

**Figure 1.**
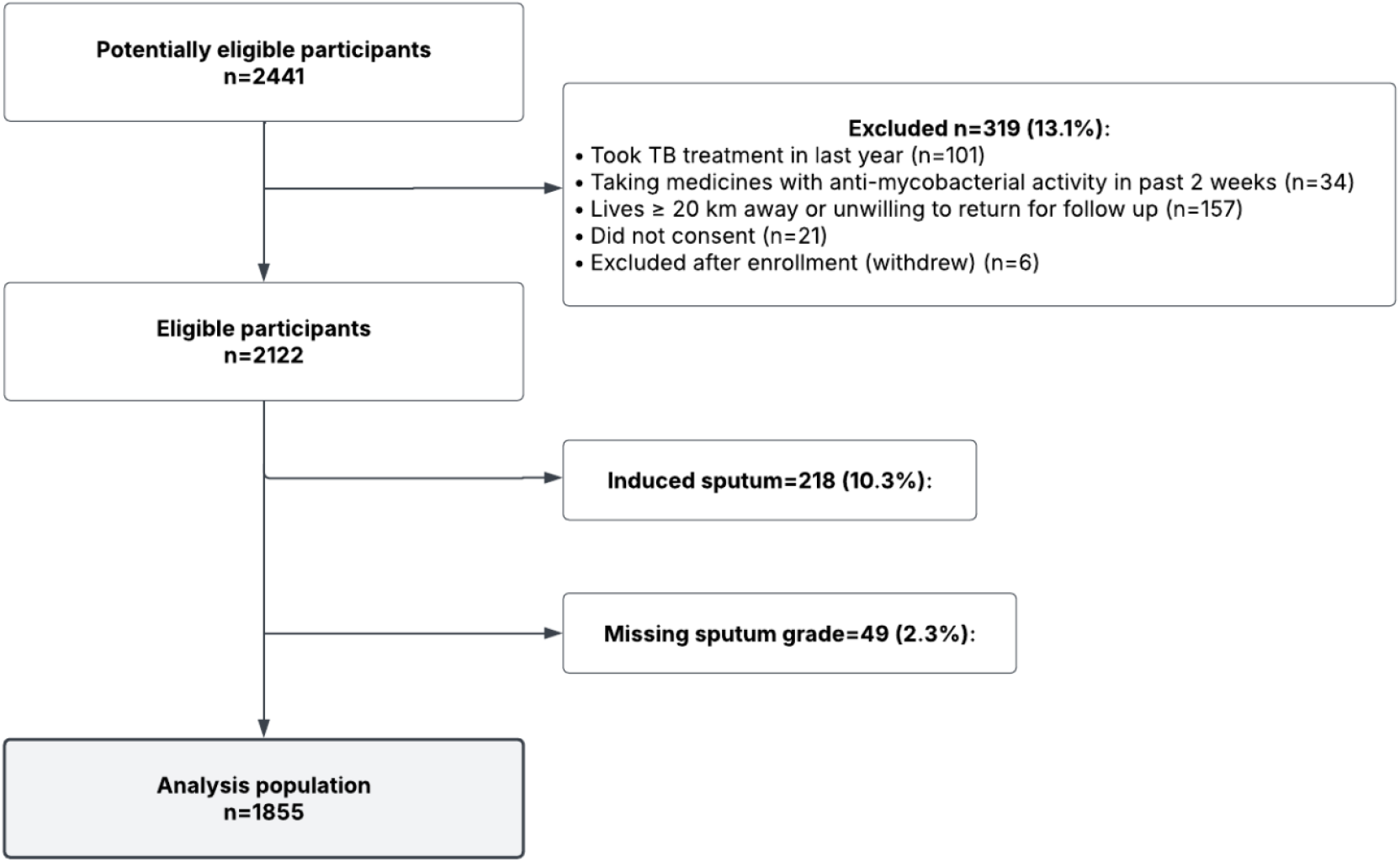
Participant flow diagram.

Among 1,855 participants, median age was 39 years (interquartile [IQR] range 29 – 52), 798 (43.0%) were female, 348 (18.9%) were living with HIV, 225 (12.1%) were living with diabetes, and 319 (17.2%) had a previous history of TB (**Table 1**). The majority of participants reported ≥2 weeks of cough (96.8%, 1795/1855). Overall, 313 (16.9%) participants tested TB-positive based on Xpert Ultra testing, while 319 (17.2%) tested TB-positive based on the MRS reference standard. Salivary sputum samples were the most common (82.6%), whereas purulent samples were the least common (1.7%) (Supplemental Table S1). Among all samples, 4.8% were blood-stained (64 salivary, 11 mucoid, 12 mucopurulent and 2 purulent samples) (**Figure 1**).

**Table 1.**
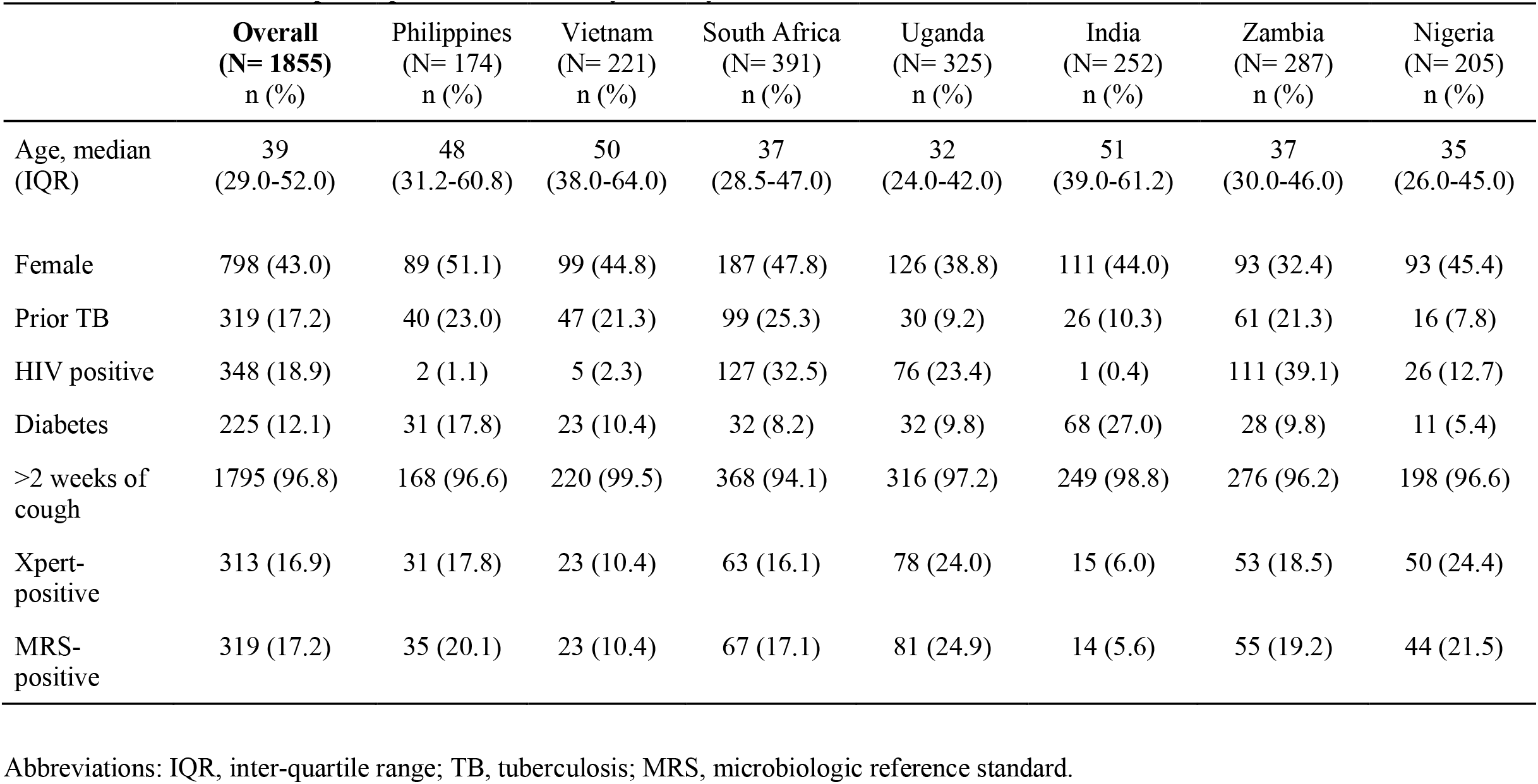
Characteristics of participants overall and by country.

### Xpert Ultra positivity

Among those with valid results, the proportion of participants with positive sputum Xpert Ultra results was 17.1% (313/1832) overall (**Figure 2 and Table S2**). Three sputum grades showed Xpert Ultra positivity close to or above 20% (mucoid 19.8% [34/172]; mucopurulent 21.7% [25/115]; and purulent 31.3% [10/32]). Xpert Ultra positivity was somewhat lower among salivary (16.1% [244/1513]) and blood-stained samples (18.2% [16/88]). In the unadjusted analysis, purulent sputum was associated with positive Xpert Ultra results (OR 2.18, 95% CI 1.02-4.67, P = 0.04) **(Table 2)**. However, after adjusting for country of enrollment, age group, sex, HIV status, abnormal chest x-ray, fever in past 30 days, and weight loss in past 30 days, there was no significant association between any sputum grade and positive Xpert Ultra results **(Table 2)**. Further, when comparing salivary vs. non-salivary sputum samples that tested positive by Xpert Ultra, there was no significant difference in the distribution of Xpert Ultra semi-quantitative grade (**Table S3**).

**Figure 2.**
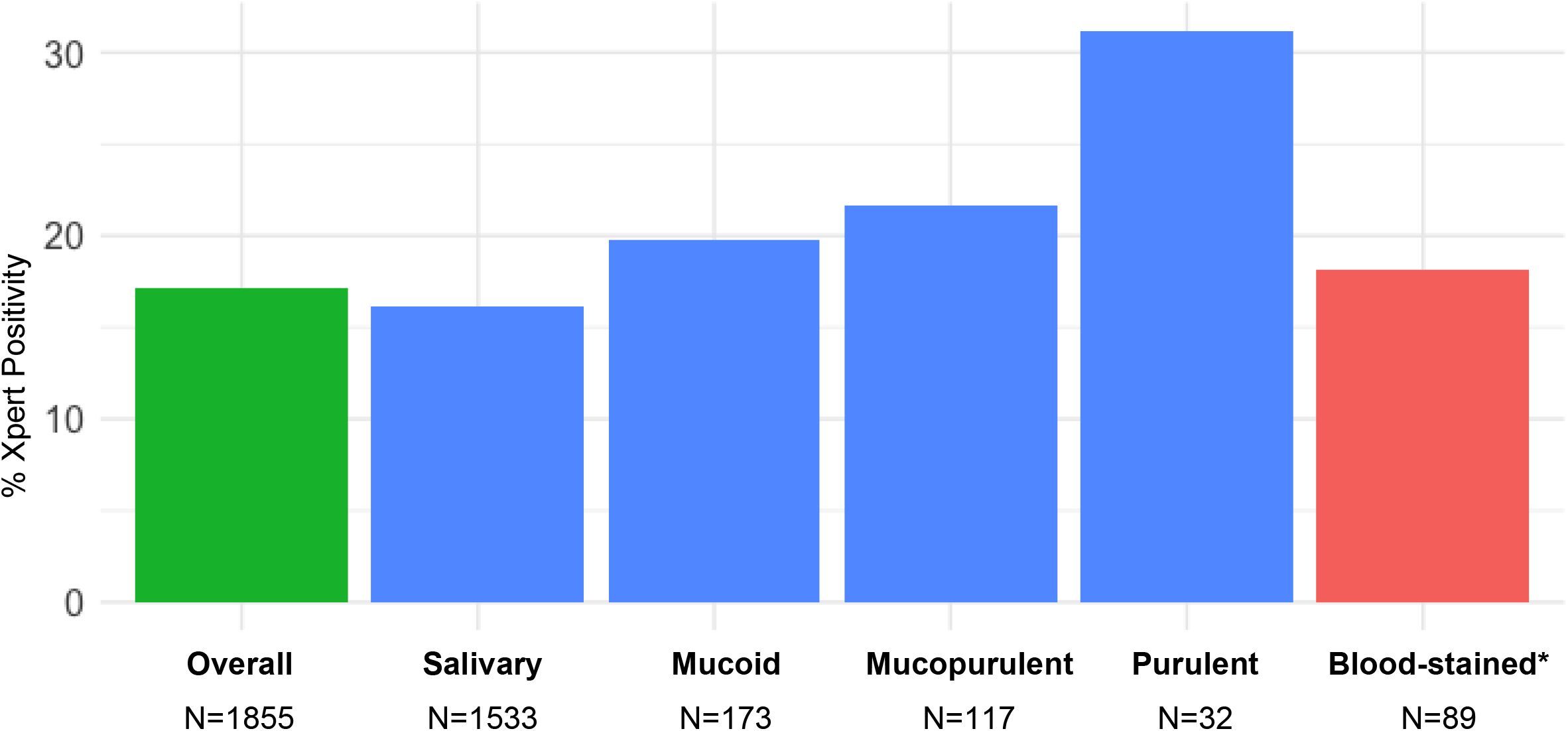
Positive Xpert Ultra results by sputum grade, N=1855. *Any sputum grade category can be blood-stained

**Table 2.** Association between Xpert Ultra positivity and sputum grade, N=1832*.

| Sputum Grade | Unadjusted Model |  | Adjusted Model** |  |
| --- | --- | --- | --- | --- |
|  | OR (95% CI) | P-value | aOR (95% CI) | P-value |
| Salivary | Ref | -- | Ref | -- |
| Mucoid | 1.22 (0.82, 1.81) | 0.32 | 1.05 (0.64, 1.72) | 0.86 |
| Mucopurulent | 1.44 (0.92, 2.26) | 0.11 | 1.08 (0.63, 1.87) | 0.77 |
| Purulent | 2.18 (1.02, 4.67) | 0.04 | 0.77 (0.30, 1.99) | 0.59 |

Notes: \* Excludes n=23 with non-actionable Xpert Ultra results \*\* Adjusted for age group, sex, country of enrollment, HIV status, chest x-ray (normal/abnormal), fever in past 30 days (yes/no), and weight loss in past 30 days (yes/no)

### Diagnostic accuracy

When compared to the MRS, the sensitivity of sputum Xpert Ultra testing was 89.6% (95% CI: 85.6-92.7, 283/316) and specificity was 98.7% (95% CI: 98.0-99.2, 1392/1410) (**Figure 3** and **Table S4**). Sensitivity was highest for purulent (100%, 95% CI: 69.2-100, 10/10) sputum samples but was high across sputum grades (salivary: 89.1% [95% CI: 84.6-92.7, 221/248], mucoid: 91.4% [76.9-98.2, 32/35], and mucopurulent: 87.0% [66.4-97.2, 20/23]). There was no significant difference in sensitivity for salivary samples vs. any other sputum grade. Specificity of sputum Xpert Ultra testing was consistently high (>98%) across all categories. However, specificity was lower for salivary samples (98.7%, 95% CI 98.0-99.2) than for mucoid (100%, 95% CI 97.0-100) and purulent (100%, 95% CI 84.6-100) samples (p<0.022 for both comparisons) (**Table S4**). Blood-stained samples also had high sensitivity (93.8%, 95% CI: 69.8-99.8, 15/16) and specificity (100%, 94.5-100, 65/65).

**Figure 3.**
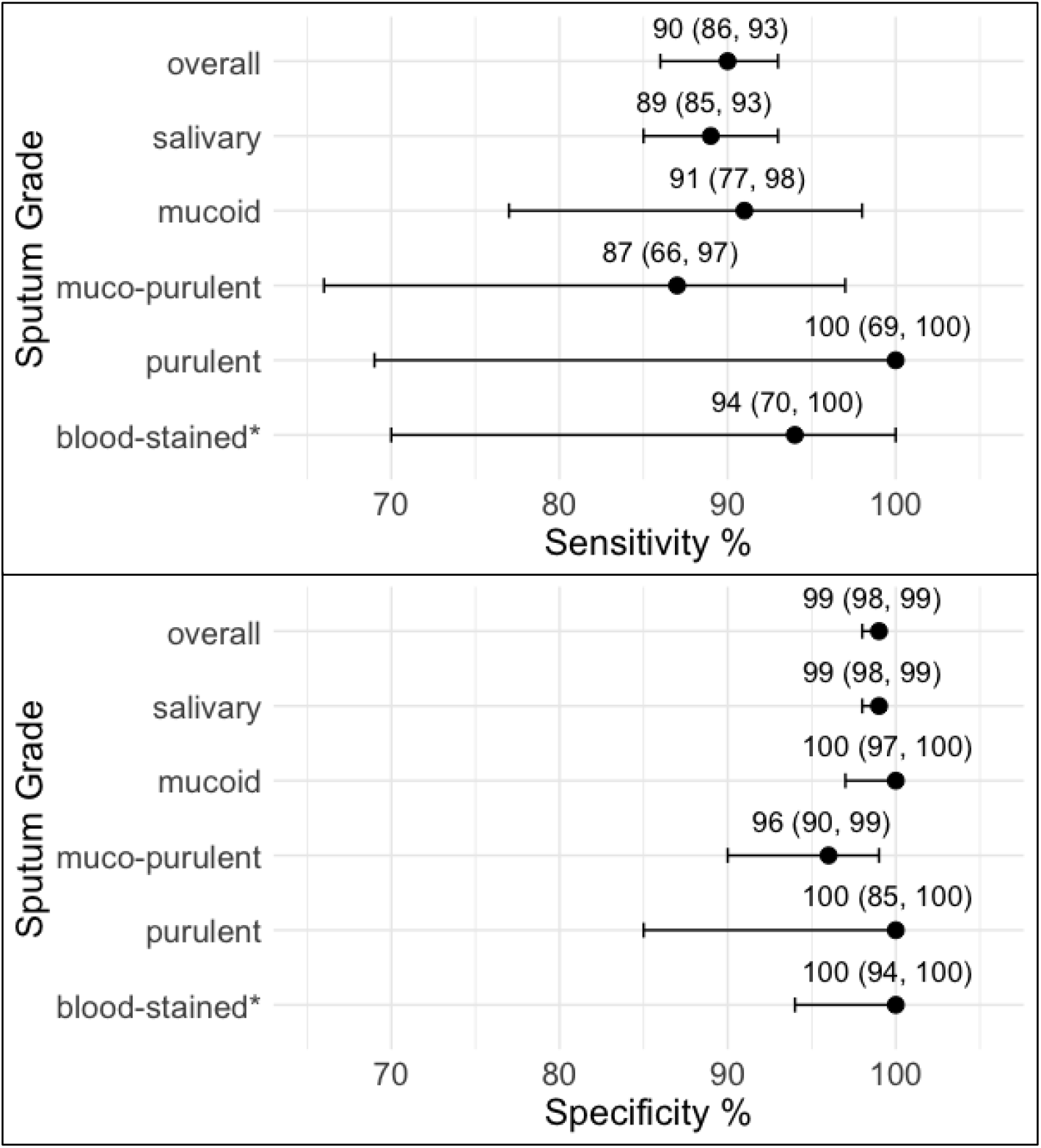
Diagnostic accuracy of Xpert Ultra in comparison to MRS, by sputum grade. *Part of the sputum samples of each grade contained blood; 64 salivary, 11 mucoid, 12 mucopurulent and 2 purulent samples were blood-stained.

#### Sensitivity analyses and robustness checks

Salivary samples were more common in females compared to males (aOR 0.74; 95% CI 0.56-0.97; p=0.03) and participants with abnormal CXR (aOR 1.41; 95% CI 1.07-1.85; p=0.01) (**Table S5**). There were also differences in frequency of salivary specimen by country of enrollment. When separating sub-categories of salivary samples, sensitivity was 100% (95% CI 29.2-100) for MRS-positive salivary-purulent samples (n=3), 89.5% (95% CI 84.7-93.2) for muco-salivary samples (n=980) and 84.6% (95% CI 65.1-95.6) for salivary samples (n=26) (**Table S6**). The sensitivity of salivary-purulent (n=3) was statistically significantly different from salivary only (n=202) specimens (p=0.048) but there were no other differences found. Xpert SPC Ct value was lower for salivary samples than for other sputum grades (**Table S7**). Compared to salivary specimens, time to culture positivity was shorter for mucoid specimens but similar for other sputum grades (**Table S8**).

## DISCUSSION

In this multi-country evaluation, macroscopic sputum quality was not associated with Xpert Ultra positivity or diagnostic sensitivity. Among 1,855 participants enrolled from seven high-burden countries, Xpert Ultra demonstrated high sensitivity and specificity across all sputum grades, including salivary specimens, which accounted for more than 80% of samples and 78% of Xpert Ultra-positive results. Critically, sensitivity on salivary sputum was approximately 90%, consistent with recent evidence from Vargas et al., who reported 90.5% sensitivity for saliva-based Xpert Ultra testing in Colombia.^20^ Although specificity varied modestly across sputum grades, differences were small in absolute terms and unlikely to be clinically meaningful. These findings indicate that the macroscopic appearance of sputum does not meaningfully influence Xpert Ultra diagnostic performance.

Our findings are consistent with prior studies in Brazil,^13^ Botswana,^14^ Ethiopia,^15^ Uganda,^16^ Vietnam,^17^ and Egypt^18^ that reported similar or higher Xpert positivity in salivary specimens, and contrast with studies in Eswatini,^10^ Fiji,^11^ and Kenya^12^ that reported modestly higher positivity or sensitivity in purulent sputum. Notably, the latter studies used the first-generation Xpert MTB/RIF assay rather than Xpert Ultra. Xpert Ultra incorporates two multicopy gene targets and has a substantially lower limit of detection than its predecessor,^3,21^ which likely allows reliable detection even in specimens with lower bacillary burden such as salivary sputum. Specimen characteristics that were important for smear microscopy or the original Xpert MTB/RIF platform are less relevant for the current Xpert Ultra assay. Our findings also help clarify conflicting prior evidence on blood-stained sputum: despite concern for PCR inhibition, blood-stained specimens in our study demonstrated high sensitivity and specificity comparable to non-blood-stained sputum of all grades, consistent with the broader pattern of performance stability across sputum types.

The heterogeneity of prior findings is further explained by inconsistent sputum grading systems, with studies using varying definitions and category structures that complicate cross-study comparisons.^22^ To our knowledge, ours is the largest and most geographically diverse evaluation addressing this issue to date, enrolling participants across sub-Saharan Africa, South Asia, and Southeast Asia, where prior studies have predominantly been single-country, single-site analyses with substantially smaller sample sizes. Our study is also the first to apply a standardized grading protocol, with written definitions, photographic reference standards, and periodic refresher training, across multiple countries and laboratories. Our use of a culture-based MRS from two specimens per participant further enables more robust sensitivity and specificity estimation than prior studies relying on smear microscopy or composite clinical criteria. Taken together, these features position our findings as the most comprehensive and directly applicable evidence for informing laboratory guidelines on sputum acceptance criteria for Xpert Ultra.

Accepting all expectorated sputum regardless of macroscopic appearance has meaningful operational implications. Rejection of salivary sputum may disproportionately exclude groups already facing diagnostic barriers, including PLHIV, individuals with early-stage disease, and those unable to produce dense expectorated specimens, without improving diagnostic yield.^23^ Eliminating quality-based rejection criteria could reduce diagnostic delays, decrease repeat clinic visits, and simplify laboratory workflows by removing a subjective, labor-intensive grading step prone to inter-operator variability.^24,25^ In high-burden settings with limited laboratory capacity, these gains could translate into earlier treatment initiation and improved outcomes along the TB diagnostic cascade.

Strengths of our study include its large sample size, multi-country scope, and standardized laboratory and grading procedures applied across all sites. The inclusion of participants from diverse high-burden countries supports the external validity of our findings. However, our study should be interpreted considering some limitations. Macroscopic sputum grading is inherently subjective. Although research staff were trained and standard definitions applied (see **Supplement**), inter-observer variation may have led to some misclassification. In addition, some sputum categories, notably purulent and blood-stained specimens, were relatively uncommon, resulting in wide confidence intervals for sensitivity estimates for those groups.

## CONCLUSIONS

Across seven high-burden countries, sputum quality based on macroscopic appearance was not associated with Xpert Ultra positivity or diagnostic accuracy. Xpert Ultra testing demonstrated high diagnostic performance across all sputum grades, including salivary samples, which accounted for 78% of all Xpert detected TB-positive results. Specificity differed modestly across grades but remained high throughout, and differences were small in absolute terms. These findings support Xpert Ultra testing of all expectorated sputum regardless of visual appearance. Where feasible, patients should be coached to produce higher-quality specimens, but salivary sputum is diagnostically useful and should not be rejected. Revising laboratory guidelines to remove sputum quality-based rejection criteria could meaningfully improve case detection, reduce diagnostic delays, and strengthen TB programs globally.

## Data Availability

All data produced in the present study are available upon reasonable request to the authors

## Acknowledgments

We gratefully acknowledge the individuals who participated in this study and extend our sincere thanks to the administrative teams, laboratory- and clinical staff at the participating health centers for their support and dedication. This work would not have been possible without their partnership and collaboration. The contents are the sole responsibility of the authors and do not necessarily reflect the views of NIH, the US Government, or consortium collaborators or members.

