## Supplement for "Impact of sputum quality on Xpert MTB/RIF Ultra test results for tuberculosis: A multi-country study"

### SUPPLEMENTAL TABLES AND METHODS

**Table S1. Sputum grades overall and by country, N=1855**

| Sputum Grade, n (%) | Overall (N= 1855) | Philippines (N= 174) | Vietnam (N= 221) | South Africa (N= 391) | Uganda (N= 325) | India (N= 252) | Zambia (N= 287) | Nigeria (N= 205) |
|---|---|---|---|---|---|---|---|---|
| Salivary | 1533 (82.6) | 157 (90.2) | 211 (95.5) | 342 (87.5) | 251 (77.2) | 187 (74.2) | 230 (80.1) | 155 (75.6) |
| Mucoid | 173 (9.3) | 0 (0.0) | 4 (1.8) | 39 (10.0) | 48 (14.8) | 35 (13.9) | 26 (9.1) | 21 (10.2) |
| Mucopurulent | 117 (6.3) | 17 (9.8) | 4 (1.8) | 8 (2.0) | 24 (7.4) | 28 (11.1) | 31 (10.8) | 5 (2.4) |
| Purulent | 32 (1.7) | 0 (0.0) | 2 (0.9) | 2 (0.5) | 2 (0.6) | 2 (0.8) | 0 (0) | 24 (11.7) |
| Blood-Stained* | 89 (4.8) | 12 (6.9) | 0 (0.0) | 25 (6.4) | 13 (4.0) | 12 (4.8) | 16 (5.6) | 11 (5.4) |

*Part of the sputum samples of each grade contained blood; 64 salivary, 11 mucoid, 12 mucopurulent and 2 purulent samples were blood-stained.

**Table S2. Proportion with positive Xpert Ultra results by sputum grade, N=1832***

| Sputum Grade | Sputum Xpert Ultra positive, % (95% CI) [n/N] |
|---|---|
| Overall | 17.1 (15.4, 18.9) [313/1832) |
| Salivary | 16.1 (14.3, 18.1) [244/1513] |
| Mucoid | 19.8 (14.1, 26.5) [34/172] |
| Mucopurulent | 21.7 (14.6, 30.4) [25/115] |
| Purulent | 31.3 (16.1, 50.0) [10/32] |
| Blood-Stained** | 18.2 (10.8, 27.8) [16/88] |

Notes:
*Excludes n=23 with non-actionable Xpert Ultra results
**Part of the sputum samples of each grade contained blood; 64 salivary, 11 mucoid, 12 mucopurulent and 2 purulent samples were blood-stained.

**Table S3. Distribution of Xpert Ultra semi-quantitative grades, N=313**

| Xpert Ultra semi-quantitative grade | Salivary (N = 244), n (%) | Non-salivary* (N = 69), n (%) | p-value |
|---|---|---|---|
| Trace | 14 (5.7) | 0 (0.0) | p = 0.07 |
| Very Low | 21 (8.6) | 7 (10.1) | |
| Low | 60 (24.6) | 12 (17.4) | |
| Medium | 56 (23.0) | 13 (18.8) | |
| High | 93 (38.1) | 37 (53.6) | |

*Includes mucoid, mucopurulent, and purulent sputum samples

**Table S4. Diagnostic accuracy of Xpert Ultra in comparison to MRS, by sputum grade, N=1726***

| | Sensitivity | | | Specificity | | |
|---|---|---|---|---|---|---|
| | % (95% CI) [n/N] | Difference, % (95% CI)** | p-value | % (95% CI) [n/N] | Difference, % (95% CI)** | p-value |
| Overall | 89.6 (85.6, 92.7) [283/316] | | | 98.7 (98.0, 99.2) [1392/1410] | | |
| Salivary | 89.1 (84.6, 92.7) [221/248] | Ref | | 98.7 (97.9, 99.3) [1167/1182] | Ref | |
| Mucoid | 91.4 (76.9, 98.2) [32/35] | 2.3 (-10.9, 14.0) | 0.81 | 100 (97.0, 100) [121/121] | 1.3 (0.4, 2.0) | 0.002 |
| Muco-purulent | 87.0 (66.4, 97.2) [20/23] | -2.2 (-16.8, 9.1) | 0.56 | 96.5 (90.0, 99.3) [82/85] | -2.3 (-6.3, 2.1) | 0.32 |
| Purulent | 100 (69.2, 100) [10/10] | 10.9 (-6.5, 3.0) | 0.47 | 100 (84.6, 100) [22/22] | 1.3 (0.2, 2.6) | 0.022 |

*Excludes n=23 with non-actionable Xpert results and n=106 with indeterminate MRS results
**95% CI and p-value adjusted for age, sex, country of enrollment, HIV status, chest x-ray (normal/abnormal), fever in past 30 days (yes/no), and weight loss in past 30 days (yes/no)

**Table S5. Factors associated with salivary sputum production, N=1855**

| | Unadjusted (bivariate) models | | Adjusted model | |
|---|---|---|---|---|
| | **OR (95% CI)** | **p-value** | **aOR (95% CI)** | **p-value** |
| Age, years | 1.01 (1.00, 1.01) | 0.17 | — | — |
| Female sex | 0.67 (0.52, 0.86) | 0.002 | 0.74 (0.56, 0.97) | 0.029 |
| *Country of enrollment* | | | — | |
| Philippines | Ref | | Ref | |
| Vietnam | 0.44 (0.20, 0.98) | 0.045 | 0.46 (0.20, 1.03) | 0.06 |
| South Africa | 1.32 (0.74, 2.37) | 0.35 | 1.37 (0.74, 2.52) | 0.32 |
| Uganda | 2.72 (1.55, 4.78) | <0.001 | 2.33 (1.29, 4.23) | 0.005 |
| India | 3.21 (1.81, 5.70) | <0.001 | 3.20 (1.77, 5.76) | <0.001 |
| Zambia | 2.29 (1.28, 4.08) | 0.005 | 1.88 (1.02, 3.46) | 0.042 |
| Nigeria | 2.98 (1.65, 5.39) | <0.001 | 2.51 (1.25, 5.02) | 0.010 |
| Positive HIV status | 1.44 (1.08, 1.92) | 0.014 | 1.39 (1.00, 1.94) | 0.052 |
| Positive diabetes status | 1.11 (0.77, 1.58) | 0.58 | — | — |
| BMI <18.5 | 1.67 (1.27, 2.20) | <0.001 | 1.12 (0.79, 1.60) | 0.51 |
| MUAC <220 | 1.88 (1.44, 2.46) | <0.001 | 1.29 (0.86, 1.96) | 0.22 |
| Anemia (defined by conjunctival pallor) | 1.00 (0.55, 1.80) | 0.99 | — | — |
| Cough ≥2 weeks | 0.93 (0.48, 1.82) | 0.84 | — | — |
| Fever | 1.42 (1.12, 1.81) | 0.004 | 0.91 (0.69, 1.21) | 0.52 |
| Chest pain | 1.13 (0.89, 1.44) | 0.33 | — | — |
| Difficulty breathing | 1.45 (1.14, 1.85) | 0.003 | 1.25 (0.96, 1.64) | 0.10 |
| Night sweats | 1.01 (0.79, 1.29) | 0.94 | — | — |
| Weight loss | 1.91 (1.49, 2.45) | <0.001 | 1.33 (1.00, 1.77) | 0.052 |
| Abnormal chest x-ray | 1.42 (1.10, 1.83) | 0.006 | 1.41 (1.07, 1.85) | 0.014 |
| History of TB | 1.30 (0.96, 1.76) | 0.09 | — | — |

Abbreviations: OR, odds ratio; aOR, adjusted odds ratio; BMI, body mass index; MUAC, mid upper arm circumference

**Table S6. Diagnostic accuracy of Xpert Ultra in comparison to MRS, by detailed sputum grade, N=1726***

| | Sensitivity | | | Specificity | | |
|---|---|---|---|---|---|---|
| | % (95% CI) [n/N] | Difference, % (95% CI)** | p-value | % (95% CI) [n/N] | Difference, % (95% CI)** | p-value |
| Salivary (N=202) | 84.6 (65.1, 95.6) [22/26] | Ref | | 98.9 (96.0, 99.9) [174/176] | Ref | |
| Mucosalivary (N=1199) | 89.5 (84.7, 93.2) [968/980] | 4.9 (-8.7, 17.8) | 0.50 | 98.8 (97.9, 99.4) [968/980] | -0.1 (-1.5, 2.1) | 0.77 |
| Salivary-purulent (N=29) | 100 (29.2, 100) [3/3] | 15.4 (0.2, 47.2) | 0.048 | 96.2 (80.4, 99.9) [25/26] | -2.7 (-8.8, 7.2) | 0.85 |
| Mucoid (N=156) | 91.4 (76.9, 98.2) [32/35] | 6.8 (-12.2, 23.5) | 0.54 | 100 (97.0, 100) [121/121] | 1.1 (-0.3, 3.1) | 0.11 |
| Muco-purulent (N=108) | 87.0 (66.4, 97.2) [20/23] | 2.3 (-17.6, 18.9) | 0.95 | 96.5 (90.0, 99.3) [82/85] | -2.4 (-6.3, 2.5) | 0.39 |
| Purulent (N=32) | 100 (69.2, 100) [10/10] | 15.4 (-10.3, 14.6) | 0.73 | 100 (84.6, 100) [22/22] | 1.1 (-0.3, 3.5) | 0.09 |

*Excludes n=23 with non-actionable Xpert results and n=106 with indeterminate MRS results

**95% CIs and p-values adjusted for age, sex, country of enrollment, HIV status, chest x-ray (normal/abnormal), fever in past 30 days (yes/no), and weight loss in past 30 days (yes/no)

**Table S7. Association between Xpert Ultra SPC Ct value and sputum grade. Coefficient interpretable per 5 units SPC Ct change, N=1726.**

| Sputum Grade | Unadjusted Model | | Adjusted Model* | |
|---|---|---|---|---|
| | Coef (95% CI) | P-value | Coef (95% CI) | P-value |
| Salivary | Ref | -- | Ref | -- |
| Mucoid | 0.07 (0.01, 13.8) | 0.028 | 0.05 (-0.01, 0.12) | 0.13 |
| Mucopurulent | 0.14 (0.07, 22.2) | <0.001 | 0.12 (0.04, 0.19) | 0.003 |
| Purulent | 0.38 (0.24, 0.53) | <0.001 | 0.22 (0.08, 0.37) | 0.003 |

* Adjusted for age group, sex, country of enrollment, HIV status, chest x-ray (normal/abnormal), fever in past 30 days (yes/no), and weight loss in past 30 days (yes/no)

**Table S8. Association between time to culture positivity (in hours) and sputum grade, N=302.**

| Sputum Grade | Unadjusted Model | | Adjusted Model* | |
|---|---|---|---|---|
| | Coef (95% CI) | P-value | Coef (95% CI) | P-value |
| Salivary | Ref | -- | Ref | -- |
| Mucoid | -84.4 (-152.5, -16.3) | 0.015 | -70.1 (-137.6, -2.6) | 0.042 |
| Mucopurulent | -14.1 (-95.8, 67.6) | 0.74 | -13.1 (-94.8, 68.6) | 0.75 |
| Purulent | 3.8 (-114.5, 122.2) | 0.95 | -34.7 (-154.7, 85.3) | 0.57 |

* Adjusted for age group, sex, country of enrollment, HIV status, chest x-ray (normal/abnormal), fever in past 30 days (yes/no), and weight loss in past 30 days (yes/no)

### Sputum Grading Methods

Sputum quality was assessed for all study participants using a standardized visual grading protocol applied either immediately after collection by health center staff or upon receipt at the laboratory. Grading was performed by trained medical technicians, laboratory technicians, or healthcare workers depending on site-specific workflows.

To assess sample quality, the grader held the sealed specimen container up to a light source and visually inspected the sample through the bottom of the container. Tilting the container to observe how the sample collected against the interior surface aided classification. Using standardized reference photographs and written descriptions, the grader assigned each sample to one of six mutually exclusive appearance categories: salivary (thin, watery, clear, non-viscous; saliva only), mucoid (viscous, clear or white; mucus only), purulent (thick, opaque, yellow or green, possibly stringy; purulent material only), mucosalivary (any mix of saliva and mucus, regardless of proportion), salivary-purulent (any mix of saliva and purulent material, with or without mucus, regardless of proportion), or mucopurulent (any mix of mucus and purulent material without saliva, regardless of proportion). Graders were instructed to assign the best-fitting category even when classification was uncertain. Additionally, each sample was independently assessed for the presence of blood-staining (yes/no), which could be recorded alongside any of the six primary appearance categories.

Sputum quality classifications were entered into REDCap via a dedicated study tablet at the time of sample collection or laboratory receipt.

Study definitions of appearance categories are defined below:

a. **Salivary**
    i. Select if entire sample is saliva only
    ii. *Texture*: thin, watery, no viscosity
    iii. *Color*: clear, may be bubbly

b. **Mucoid**
    i. Select if entire sample is Mucus only
    ii. *Texture*: viscous
    iii. *Color*: May be clear or white

c. **Purulent**
    i. Select if entire sample is purulent material only
    ii. *Texture*: thick, may be stringy
    iii. *Color*: opaque with a yellow or green tinge

d. **Mucosalivary**
    i. Select if sample includes any mix of saliva and mucus (regardless of proportion salivary or mucoid)

e. **Salivary-purulent**
    i. Select if sample includes any mix of saliva and purulent material (regardless of proportion salivary or purulent), with or without mucus

f. **Mucopurulent**
    i. Select if sample includes any mix of mucus and purulent material (regardless of proportion mucoid or purulent), with no saliva

g. **Blood-stained**
    i. Any of the above described appearance categories can be blood-stained. Select “yes” if you think the specimen is blood stained and “No” if you think specimen is not blood stained.

**Examples of Sputum Samples by Type**

| | **Sample appearance** | **Classification** |
|---|---|---|
| 1. | 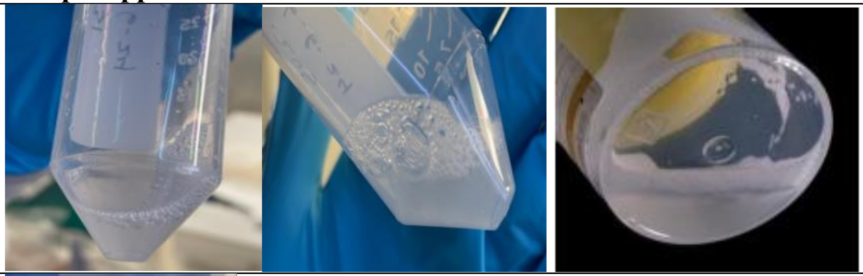 | Salivary |
| 2. | 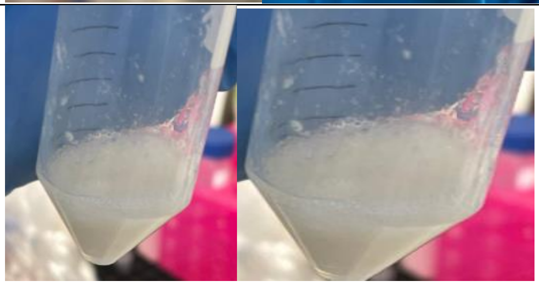 | Mucoid |
| 3. | 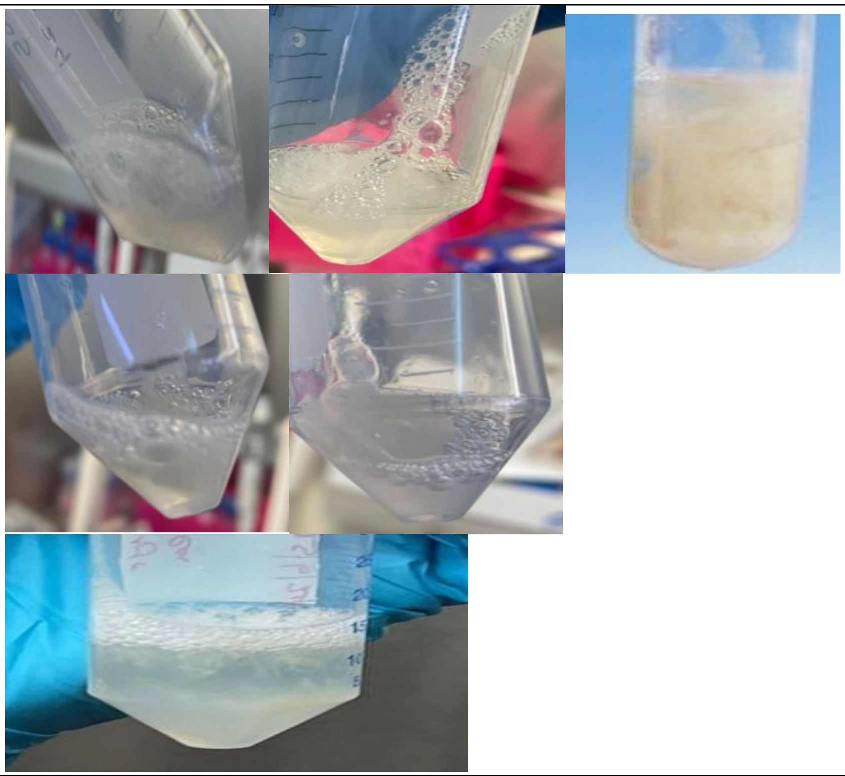 | Muco-salivary |

| | | |
|---|---|---|
| 4. | 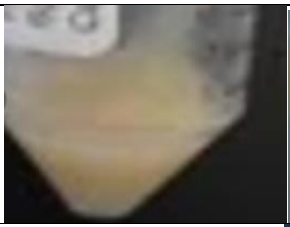 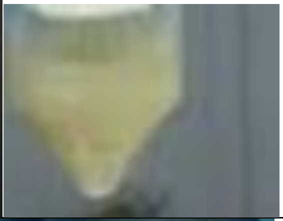 | Muco-Purulent |
| 5. | 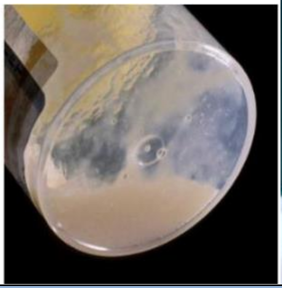 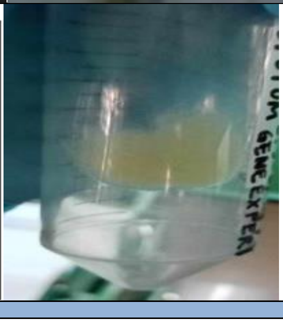 | Purulent |
| (any of the above) | 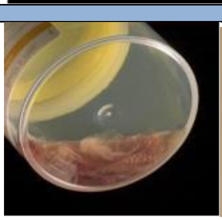 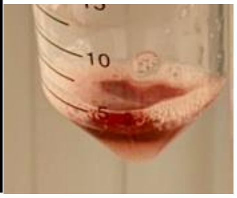  | Blood-stained |
